# Evaluating Large Language Models for Translating Multimodal Phenotype Documentations into Executable EHR Phenotyping Algorithms

**DOI:** 10.64898/2026.05.20.26353690

**Authors:** Chao Yan, Yi Xin, Wu-Chen Su, Srushti Gangireddy, Shravani Durbhakula, Stephen P. Bruehl, Alyson L. Dickson, Lang Li, QiPing Feng, Bradley A. Malin, Tyler Derr, Wei-Qi Wei

## Abstract

Research applications of electronic health record (EHR) phenotypes require translating clinical definitions into executable EHR database queries, a labor-intensive process. We evaluated two frontier large language models across five phenotypes and three documentation modalities. Both models captured high-level logic from structured text but degraded markedly with diagram-only input. Error analysis revealed seven failure categories. Documentation, rather than model capability, was the primary bottleneck, reinforcing the need for standardization and expert oversight.

## Main

The ability to characterize clinical phenotypes using electronic health records (EHRs) underpins a wide range of biomedical studies. These characterizations are often accomplished through phenotyping algorithms (or computable definitions) that identify patients’ observable traits from routine care data at scale.^1^ Phenotyping algorithms are often constructed as rule-based criteria that combine diagnoses, medications, laboratory measurements, procedures, and demographics into explicit inclusion and exclusion criteria.^2,3^ While the utility of such algorithms has been highlighted through numerous studies, the formulation of them requires manual translation into computational instructions that are tailored to site-specific conventions that account for local data models and coding systems, followed by iterative validation through extensive expert review and chart abstraction.^4^ This labor-intensive process remains a major bottleneck, limiting the portability, comparability, and reuse of cohorts across biomedical research.

A primary reason for this bottleneck is that phenotyping algorithms are rarely communicated in a computation-ready form. Rather, they are typically described and documented at a high level of abstraction intended for human readability and interpretation; these descriptions range across various modalities, such as narrative text, tables, flowcharts, and diagrams. They must then be manually translated into structured query language (SQL) to run against EHR databases. Even well-documented algorithms, such as those deposited in PheKB,^5^ often fragment key implementation details, such as index date, temporal anchors and lookback windows, threshold and unit specifications, code-set boundaries, and conditional branching across formats or leave them implicit. As a result, successful reuse depends heavily on analyst expertise, domain knowledge, and time investment. This introduces interpretive variability that can lead to inconsistent implementations and ultimately divergent cohort definitions across sites.

Large language models (LLMs) provide a potential remedy. Recent research suggests that LLMs can assist phenotyping by extracting candidate clinical criteria and expressing them in forms aligned with common data models (CDM).^6,7^ More broadly, LLMs have demonstrated strong performance in multi-step reasoning and code generation tasks,^8–10^ which motivate their use as translation engines from human-authored specifications to executable implementations. However, it remains unclear whether LLMs can faithfully convert real-world, multimodal phenotype documentation into standards-conformant, executable SQL code. Here, we systematically evaluate LLM-based translation of phenotyping algorithms into Observational Medical Outcomes Partnership (OMOP) SQL, testing how input modality shapes the correctness, completeness, and clinical fidelity of the generated queries.

In this study, we focus on two frontier LLMs, i.e., OpenAI GPT o3 and Claude Opus 4.1, and five established clinical phenotypes selected from PheKB: 1) acute kidney injury (AKI), 2) autoimmune diseases (ADs), 3) familial hypercholesterolemia (FH), 4) major adverse cardiovascular events (MACE), and 5) type 2 diabetes mellitus (T2DM). Two clinical phenotyping experts (W.C.S. and S.G.) were invited to assess generated SQL queries along seven complementary dimensions using a rating range 0-4: 1) logical and Boolean accuracy, 2) temporal constraint implementation, 3) value-set precision, 4) OMOP schema and domain correctness, 5) coding efficiency, 6) human readability, and 7) revision effort for local deployment (readiness). See **Supplementary Tables 1 and 2** for detailed prompt and expert evaluation criteria, respectively. The interrater agreement between the two clinical experts, as measured by Cohen’s kappa with quadratic weights, achieved 0.798 and 0.898 for OpenAI o3 and Claude Opus 4.1, respectively, indicating substantial agreement according to Landis and Koch criteria (See **Supplementary Table 3** for disease-level agreement scores).^11^

All generated SQL queries were executed successfully within the OMOP CDM environment after minor fixes. There are several notable findings from our evaluation. First, the input modality had a significant and consistent effect on query quality across both models (**Fig. 1a, b**). Providing only diagrams resulted in dramatically lower ratings compared to full-document or text-only scenarios, with particularly pronounced degradation in value-set precision, temporal constraint implementation, logic and Boolean accuracy, and readiness for local deployment. The first three of these dimensions all require precise, structured clinical reasoning. Second, text-only input yielded performance closely approximating full-document input for both OpenAI o3 and Claude Opus 4.1 (**Fig. 1a, b**), suggesting that the narrative prose of phenotyping documents captures the majority of information necessary for SQL generation, while visual phenotyping diagrams alone are insufficient as a standalone input modality. Third, direct comparison of the two models revealed that Claude Opus 4.1 achieved better or comparable scores in five of the seven dimensions under both full-document and text-only conditions, while OpenAI o3 outperformed Claude Opus 4.1 specifically in logical and Boolean accuracy and readiness for local deployment (**Fig. 1c, d**). Under the diagram-only condition, this gap narrowed considerably, with OpenAI o3 maintaining an advantage only in logical and Boolean accuracy (**Fig. 1e**). Fourth, performance across the seven evaluation dimensions was not uniform, with human readability, OMOP schema correctness, and coding efficiency demonstrating greater higher ratings in logical and Boolean accuracy, temporal constraint implementation, and value-set precision for autoimmune disease relative to the other four phenotypes, resulting in correspondingly higher overall ratings (**Supplementary Figs. 1**,**2**). This advantage likely reflects the structure of the underlying phenotyping document, which articulates eligibility criteria and clinical logic in explicit, well-organized pseudo-code, a level of specification absent from the documentation of the remaining diseases. Collectively, these findings indicate that the quality of LLM-generated clinical phenotyping queries is highly sensitive to input representation, with structured natural language being both necessary and largely sufficient for near-optimal performance.

**Fig. 1:**
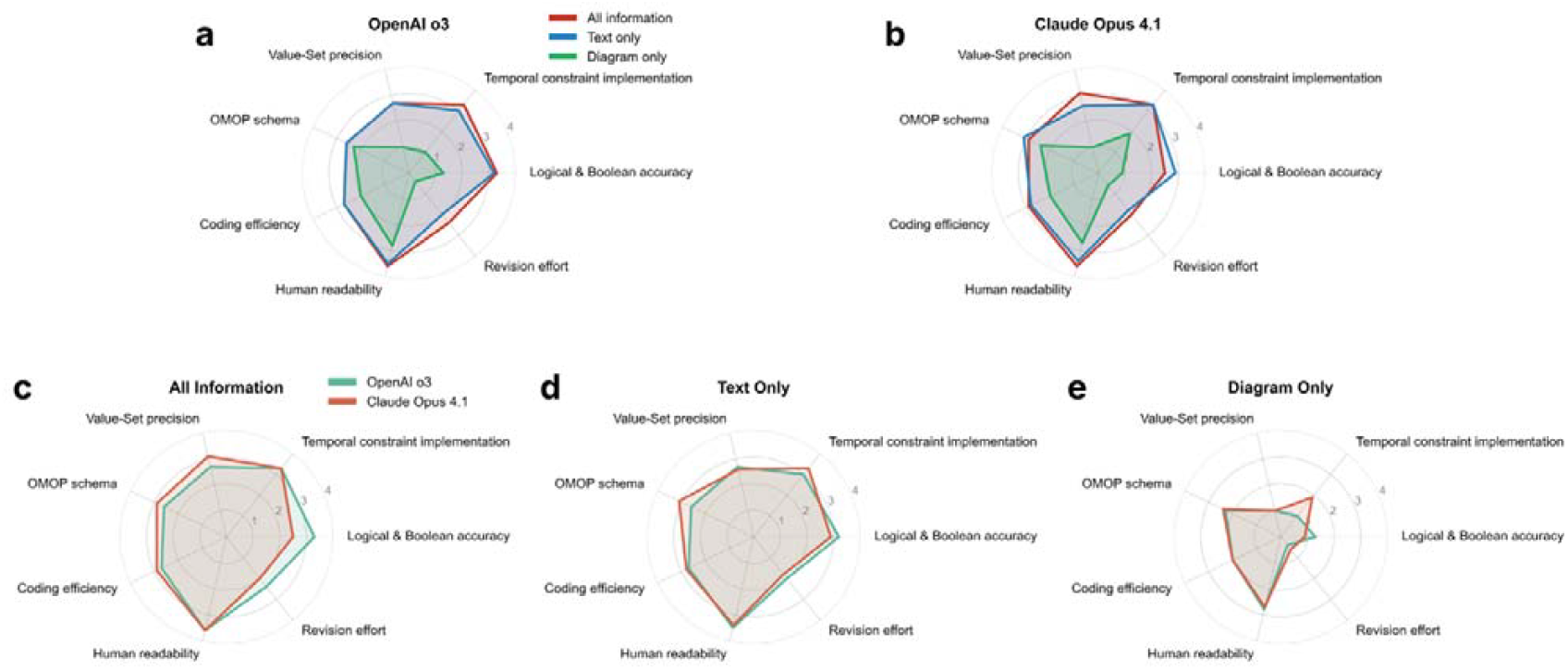
Expert evaluation of LLM-generated SQL queries. All panels show mean ratings from two experts across five clinical phenotypes. **a**, OpenAI o3. **b**, Claude Opus 4.1. **c**, LLMs provided with full phenotyping documents. **d**, LLMs provided with text narratives only. **e**, LLMs provided with diagrams only.

A qualitative error analysis of the LLM-generated SQL queries identified seven distinct error categories that collectively illuminate the failure modes of current frontier models (**Table 1**). Logical fidelity errors were among the most consequential, involving fundamental misrepresentation of phenotype structure. For example, OpenAI o3 incorrectly treated a single diagnosis or laboratory result as sufficient for autoimmune disease case ascertainment, rather than enforcing the required pattern of repeated diagnoses over time. Closely related were under-specification and over-generalization errors, in which models collapsed multi-pathway case definitions into broad OR conditions, discarding the sequencing and temporal dependencies essential to accurate cohort identification. Concept-set incompleteness represented another recurring failure, with both models omitting required medication, laboratory, or procedural concept IDs, leading in some cases (notably for AKI under diagram-only input) to near-zero code accuracy and clinically unusable output. Schema and implementation errors further compromised deployability, manifesting as placeholder table names lacking OMOP concept IDs, incorrect domain mappings, and string-based concept matching that introduced misclassification risk and reduced portability across institutional environments. Threshold and parameter errors, though subtler, were clinically significant: for example, Claude Opus 4.1 applied an incorrect AKI staging threshold (≥3.0× baseline serum creatinine instead of the specified strictly >3×), which would systematically alter cohort composition. Hallucinations constituted a particularly concerning error type, with Claude Opus 4.1 introducing unsupported clinical concepts (e.g., hallucinated autoimmune conditions and encounter-type distinctions not grounded in OMOP vocabularies), causing substantive phenotype drift. Finally, modality interpretation limitations were especially prominent under diagram-only conditions, where both models failed to translate visual specifications into correct temporal logic; in particular, crucial exclusion criteria (e.g., triglyceride thresholds and secondary cause exclusions for familial hypercholesterolemia) were systematically omitted from the generated SQL queries in the diagram-only modality.

**Table 1:** A summary of error categories and corresponding examples found in LLM-generated SQL queries, as reviewed by clinical phenotyping experts. Diseases are listed in no particular order. ADs: autoimmune disease; AKI: acute kidney injury; FH: familial hypercholesterolemia; MACE: major adverse cardiac events; T2DM: type 2 diabetes mellitus.

| Error type | Phenotype | Modality | LLM | Description |
| --- | --- | --- | --- | --- |
| Logical fidelity errors | ADs | All | o3 | Case logic incorrectly treats a single diagnosis or lab result as sufficient, instead of enforcing repeated diagnoses over time, fundamentally changing the intended phenotype meaning. |
|  | T2DM | Text | Opus 4.1 | The case pathways and threshold logic are incomplete and partially incorrect, with missing or simplified conditions and inaccurate diagnostic and lab boundary cutoffs. |
| | FH | Diagram | o3 | Core phenotype entry criteria (individuals $\geq 18$ years old with lipid panel) are missing, and category definitions are altered, resulting in logic that no longer matches the intended phenotype structure. |
| Concept-set incompleteness | FH | All | Opus 4.1 | Required concept sets are incomplete (e.g., lipid medications, HDL, secondary causes, procedures), resulting in partial phenotype coverage. |
|  | MACE | Text | o3 | Statin medication concept IDs are incorrect or incomplete, weakening medication-based exclusions and cohort validity. |
|  | AKI | Diagram | Opus 4.1 | Numerous required diagnosis and laboratory concept IDs are missing or incorrect, leading to near-zero code accuracy and unusable SQL query. |
| Schema and implementation errors | ADs | All | o3 | The SQL query relies on placeholder table names and omits actual OMOP concept IDs, making the implementation non-executable without substantial schema mapping. |
|  | AKI | Text | o3 | Required OMOP domains (e.g., observation for end-stage renal disease-related concepts) are incompletely used, and repeated full-table measurement scans reduce efficiency and deployability. |
|  | MACE | Diagram | Opus 4.1 | String-based concept matching and hard-coded schema assumptions are used instead of explicit OMOP concept joins, increasing misclassification risk and limiting portability. |
| Threshold and parameter errors | ADs | All | Opus 4.1 | Serology positivity thresholds are imprecise or overly simplistic, using generic comparisons that do not align with the phenotype specification. |
| | AKI | Text | Opus 4.1 | Incorrect threshold definition ($\geq 3.0 \times$ baseline SCr) is used instead of the specified strictly $> 3 \times$ baseline, altering AKI staging. |
| | FH | Diagram | o3 | core phenotype entry criteria such as individuals $\geq 18$ years old with lipid panel and triglyceride level $\geq 500$ mg/dL exclusion are missing. |
| Hallucinations | T2DM | All | Opus 4.1 | Encounter-type and family-history concepts are hallucinated, including unsupported inpatient/outpatient distinctions not grounded in OMOP vocabularies. |
|  | ADs | All | Opus 4.1 | Disease lists include hallucinated autoimmune conditions while omitting many required real diseases, causing major phenotype drift. |
| | AKI | Text | Opus 4.1 | The SQL query introduces a serum creatinine threshold (SCr $< 30$) not present in the phenotype specification, representing unsupported clinical logic. |
| Under-specification / over-generalization | T2DM | Text | o3 | The case definition is over-generalized by collapsing multiple required pathways into broad OR conditions, omitting sequencing, visit counts, and temporal constraints. |
|  | ADs | Text | o3 | Serology positivity logic is applied too generically, using simplified threshold handling without sufficient constraint or context, potentially over-including cases. |
| Modality interpretation limitations | MACE | Text | Opus 4.1 | Narrative temporal logic is implemented using absolute date differences rather than event-anchored windows, reflecting partial modality interpretation. |
|  | AKI | Diagram | Opus 4.1 | Diagram-based AKI block separation and recurrence timing logic are not correctly translated into the SQL query, resulting in incorrect temporal reasoning. |
| | FH | Diagram | o3 | Key exclusions (e.g., triglycerides $\geq 500$ mg/dL, secondary causes) appear only in diagrams and are omitted from the generated SQL query, indicating failure to integrate visual specifications. |

Our findings have several key implications. First, encouragingly, both models demonstrated strong performance in reconstructing high-level logic and temporal constraints when provided with full phenotyping documentation, capturing the general structure and flow of complex, multi-layered phenotype algorithms with meaningful fidelity. This is notable given that phenotype translation has historically required substantial expert effort, limiting scalability and cross-site reuse. Second, query quality was highly dependent on input modality: performance degraded markedly with diagram-only input, while text-based representations were largely sufficient. This observation suggests that current LLMs should not be treated as general-purpose interpreters of arbitrary phenotyping artifacts. Instead, their use is best supported when documentation is provided in structured natural language, and practitioners should avoid relying solely on diagram-based specifications as LLM inputs. Third, phenotypes expressed with explicit pseudo-code consistently yielded the highest-quality outputs, pointing to a promising path forward: standardizing phenotype documentation into implementation-oriented formats can substantially improve translation fidelity without requiring advances in model design. Fourth, the diversity and clinical significance of the identified error categories underscore that LLM-generated SQL queries are not yet suitable for autonomous deployment. Mandatory expert review remains essential, particularly for dimensions such as temporal constraint implementation and concept-set precision that proved most sensitive to input modality and most consequential for cohort validity. Fifth, the comparable yet complementary performance profiles of OpenAI o3 and Claude Opus 4.1 suggest that ensemble or consensus-based generation strategies, wherein outputs from multiple models are cross-validated, may offer a practical means of mitigating model-specific failure modes. Collectively, these results reframe the role of LLMs in phenotyping not as autonomous implementers, but as capable first-draft generators whose utility is gated by documentation quality and augmented by structured human oversight, a collaborative paradigm that, if adopted systematically, could meaningfully accelerate the portability and reuse of clinical phenotypes across institutions and research networks.

This study has several limitations. First, the evaluation was conducted on five phenotypes from PheKB. Although diverse, this sample may not fully capture the full spectrum of phenotyping complexity, and broader, systematically sampled libraries are needed to support generalizability. Second, only two frontier LLMs were evaluated, and given the rapid pace of model development, the relative performance profiles reported here may shift as more advanced model generations are released. Longitudinal benchmarking frameworks that can accommodate emerging models will be essential. Third, expert evaluation was performed by two raters, and although interrater agreement was substantial, the subjective nature of dimensional scoring introduces some degree of variability. Larger panels of independent reviewers with diverse institutional backgrounds would strengthen the reliability of quality assessments. Fourth, although all generated SQL queries were successfully executed within an OMOP CDM environment, this evaluation focused primarily on implementation validity and semantic fidelity rather than downstream cohort accuracy or prospective clinical deployment. Future work should incorporate cohort-level validation against gold-standard chart-reviewed datasets and multi-site execution testing to assess transportability across heterogeneous OMOP environments. Fifth, prompt engineering was not systematically optimized in this study, and it is possible that structured prompting strategies, such as few-shot examples and retrieval-augmented generation with OMOP vocabulary resources, could meaningfully improve performance, particularly for dimensions such as concept-set precision and temporal constraint implementation that proved most challenging under the current evaluation conditions.

In conclusion, LLMs can generate executable and useful first-draft phenotyping queries, but their performance depends strongly on documentation. While structured text enables accurate translation, multimodal and under-specified artifacts introduce substantial errors. These findings position LLMs as assistive tools rather than replacements for expert implementation and highlight documentation standardization as key to scalable, reproducible phenotyping.

## Methods

We selected five phenotyping algorithms from PheKB^5^, a publicly accessible, community-driven repository that aggregates phenotyping algorithms developed and validated across multiple institutions, serving as a *de facto* standard resource for sharable and reusable EHR-based phenotype definitions. The five phenotypes were 1) acute kidney injury (AKI), 2) autoimmune disease (ADs), 3) familial hypercholesterolemia (FH), 4) major adverse cardiovascular events (MACE), and 5) type 2 diabetes mellitus (T2DM). These phenotypes were selected because they collectively represent a range of clinical domains and algorithmic complexity, with documentation spanning diverse representational formats including narrative text, flowcharts, tables, and pseudo-code. For example, the AKI algorithm relies on detecting ≥50% increases in serum creatinine relative to baseline and evaluating consecutive day patterns to define AKI blocks, staging, and subtypes. The T2DM algorithm identifies cases using a combination of T2DM diagnosis codes, diabetes-related medications, and abnormal glucose or HbA1c laboratory thresholds. This diversity in logic structure and documentation style makes these algorithms well-suited benchmarks for evaluating LLM performance in generating phenotyping SQL queries.

We evaluated two frontier LLMs: OpenAI o3^12^ and Claude Opus 4.1^13^. At the time of the study, OpenAI o3 represented OpenAI’s most advanced reasoning-oriented model, while Claude Opus 4.1 was Anthropic’s highest-capacity model optimized for complex analytical tasks. Both models were selected on the basis of their demonstrated strengths in multi-step reasoning and code generation, making them strong candidates for evaluating state-of-the-art LLM performance in this domain.

We designed a four-stage prompting pipeline to generate executable OMOP-compliant SQL queries from phenotyping documentation. In the first stage (Prompt 1), the model was instructed to extract a structured pseudo-algorithm from the input document (PDF) using chain-of-thought reasoning, guided by the phenotype’s logical structure and relevant OMOP concepts. In the second stage (Prompt 2), the model was asked to verify the generated pseudo-algorithm and all its details were grounded in the provided document. In scenarios where errors were found, the model was instructed to produce a new version with the errors corrected. In the third stage (Prompt 3), the corrected pseudo-algorithm was used to generate an executable SQL query aligned with the OMOP CDM. In the fourth stage (Prompt 4), the model was instructed to review and correct the generated SQL queries when errors were found. All four prompt templates are provided in **Supplementary Table 1**.

To systematically examine the influence of input modality on generation quality, we defined three input scenarios. Scenario 1 (All Information) provided the full phenotyping document, including narrative text, tables, diagrams, and pseudo-code. Scenario 2 (Text Only) retained only the narrative text and tables, excluding diagrams and pseudo-code. Scenario 3 (Diagram Only) provided solely the visual diagrams, isolating the contribution of graphical representations. Applying all three scenarios to five phenotypes across two models yielded a total of 30 generated SQL queries. All generated SQL queries were executed within an OMOP CDM environment to assess implementation validity prior to expert evaluation.

Expert evaluation of the generated SQL queries was conducted along seven dimensions as mentioned above. Two experienced clinical phenotyping experts independently scored each query on a 0-4 scale for every dimension. Interrater reliability was quantified using Cohen’s kappa with quadratic weights. During scoring, the experts performed a deeper qualitative review of each query to identify specific errors and areas requiring correction.

We categorized the identified errors into seven categories: 1) logical fidelity errors, referring to failures to correctly interpret or implement the intended phenotype logic; 2) concept-set incompleteness, referring to missing or incomplete clinical concept IDs, value sets, or code lists (e.g., LOINC, ICD, RxNorm); 3) schema and implementation errors, referring to incorrect or suboptimal use of the OMOP CDM schema or SQL implementation details; 4) threshold and parameter errors, referring to incorrect numeric thresholds; operators, or parameter definitions; 5) hallucinations, defined as generated logic, thresholds, or conditions not present in the phenotype specification; 6) under-specification or over-generalization, characterized by oversimplification of multi-step logic, collapsed decision branches, or generalized criteria; and 7) modality interpretation limitations, describing errors from the model’s difficulty integrating information across text, diagrams, and pseudo-code.

## Acknowledgement

This work is supported in part by National Institute of Health grants K99LM014428 and U54HG012510.

## Author Contribution Statement

W.Q.W., C.Y., and Y.X. conceived and designed this study. C.Y. and Y.X. conducted data preprocessing, performed the experiments, analyzed the results, summarized the major experimental findings, and drafted the manuscript. W.C.S. and S.G. reviewed and evaluated LLM-generated phenotyping SQL codes as phenotyping experts. T.D. and B.A.M. assisted in interpreting the results and provided significant intellectual feedback. A.L.D., L.L., Q.F., S.D., S.P.B., T.D., and B.A.M. extensively revised the manuscript. W.Q.W. supervised the study. All authors participated in manuscript preparation and approved the final version.

## Ethics Declarations

There is no conflict of interest for this study.

## Code and Data Availability

All source codes and data used and produced in this study are available at https://github.com/yanchao0222/LLM_phenotyping_doc_translation

## Supplementary Information

**Supplementary Fig. 1:**
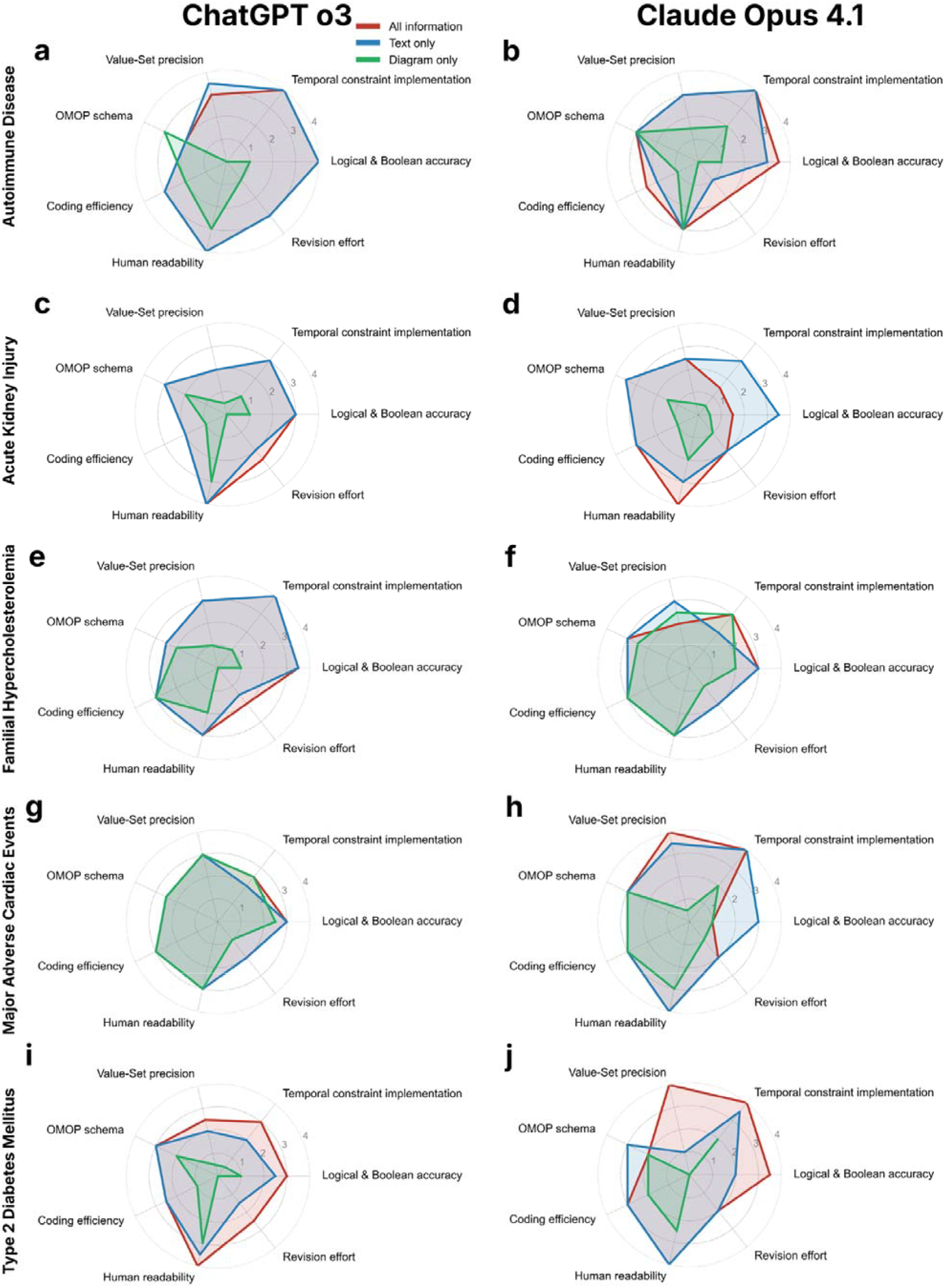
Expert evaluation of LLM-generated OMOP SQL queries. Mean ratings for seven dimensions stratified by two LLMs and five clinical phenotypes: **ab**, Autoimmune disease. **cd**, Acute kidney injury. **ef**, Familial hypercholesterolemia. **gh**, Major adverse cardiac events. **ij**, T2DM: type 2 diabetes mellitus.

**Supplementary Fig. 2:**
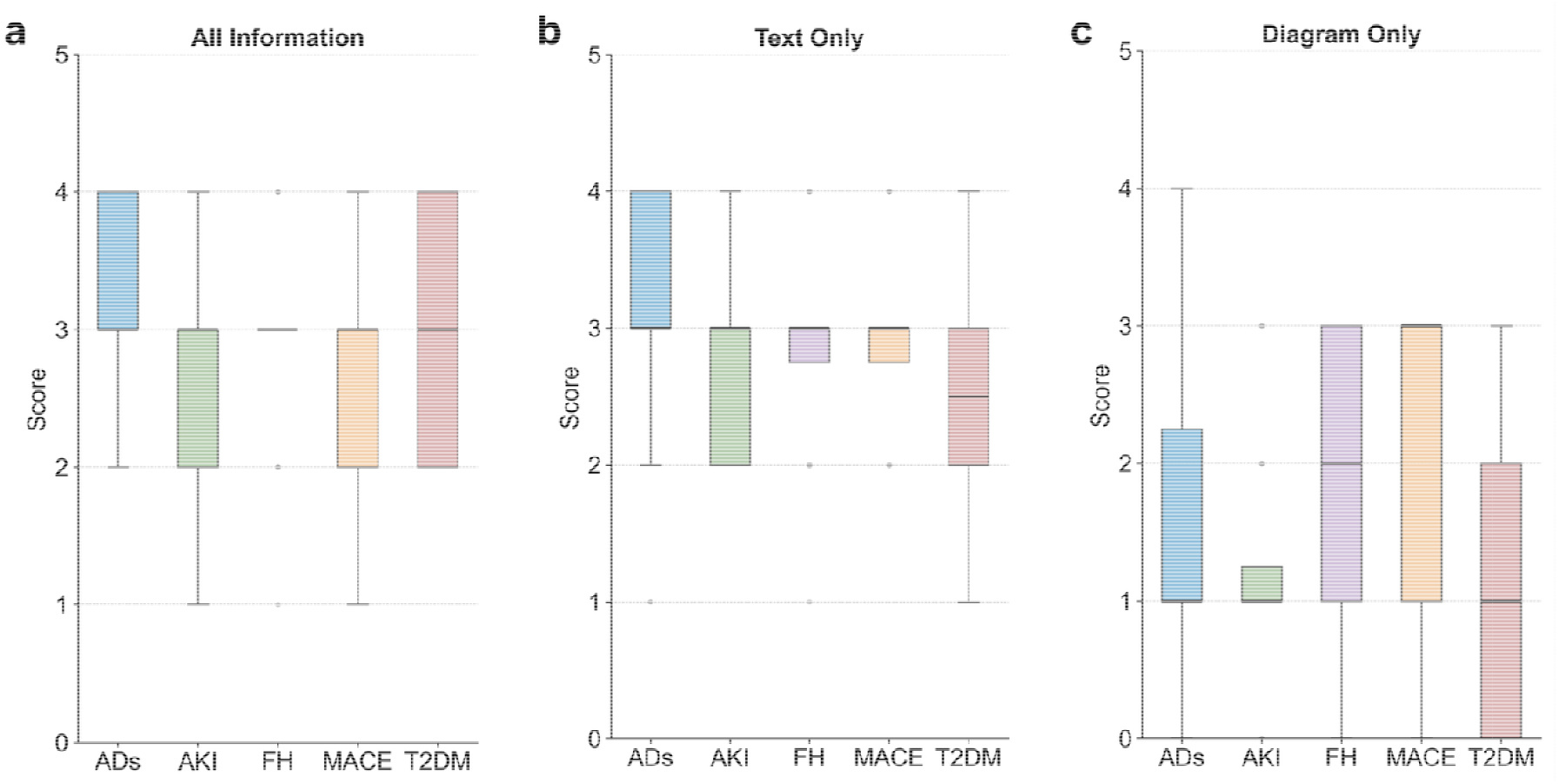
Expert evaluation of LLM-generated OMOP SQL queries across OpenAI o3 and Claude Opus 4.1 for five clinical phenotypes stratified by information settings: **a**, All information; **b**, Text only; **c**, Diagram only. ADs: autoimmune disease; AKI: acute kidney injury; FH: familial hypercholesterolemia; MACE: major adverse cardiac events; T2DM: type 2 diabetes mellitus.

**Supplementary Table 1:**
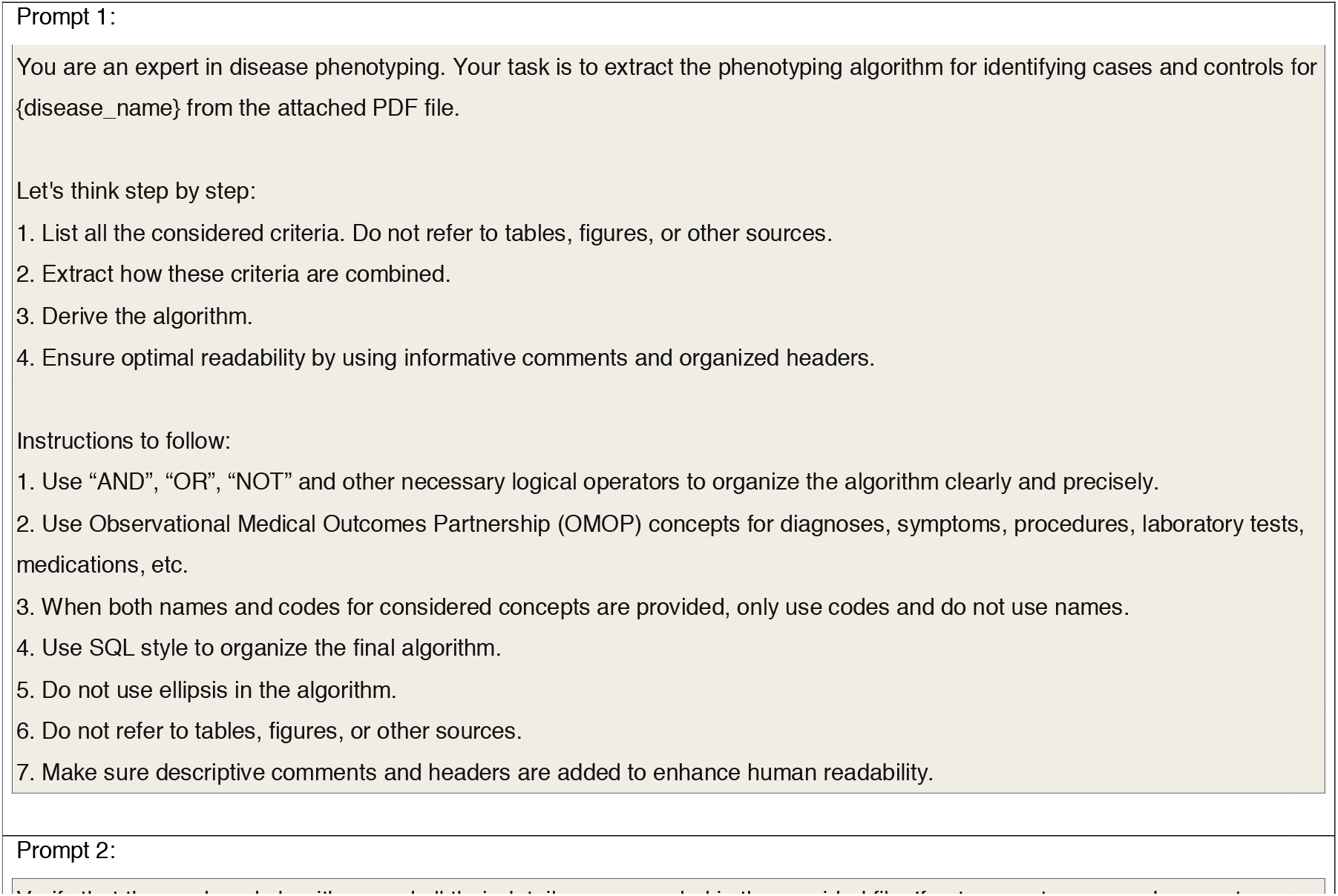
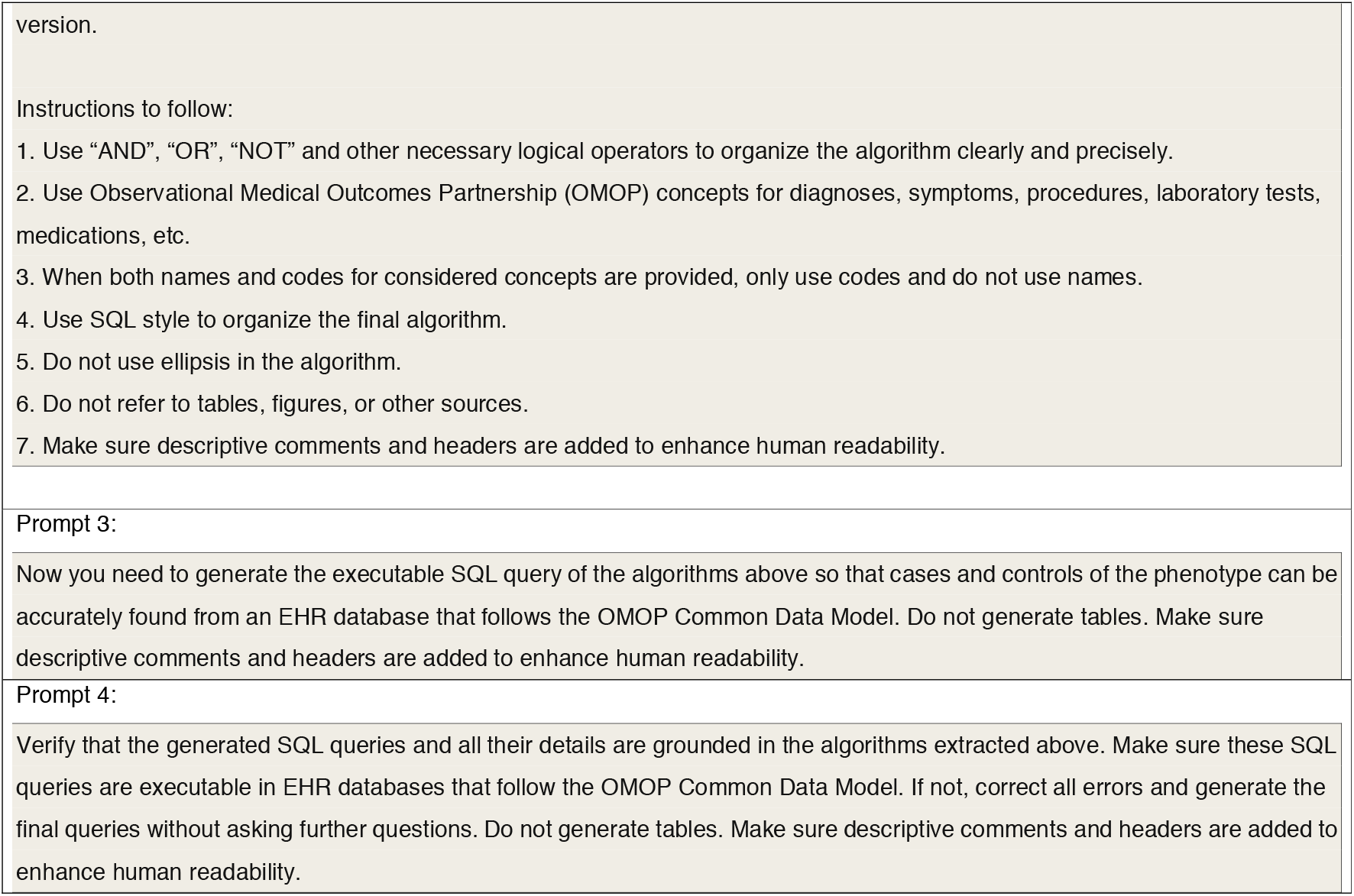
Prompt templates for stepwise LLM-based phenotype implementation. A sequential four-prompt chain decomposes the task into two phases: algorithm extraction (prompts 1&2), in which clinical logic is parsed from PDF documentation, and SQL generation (prompts 3&4), in which the extracted logic is rendered as executable OMOP-conformant queries.

**Supplementary Table 2:**
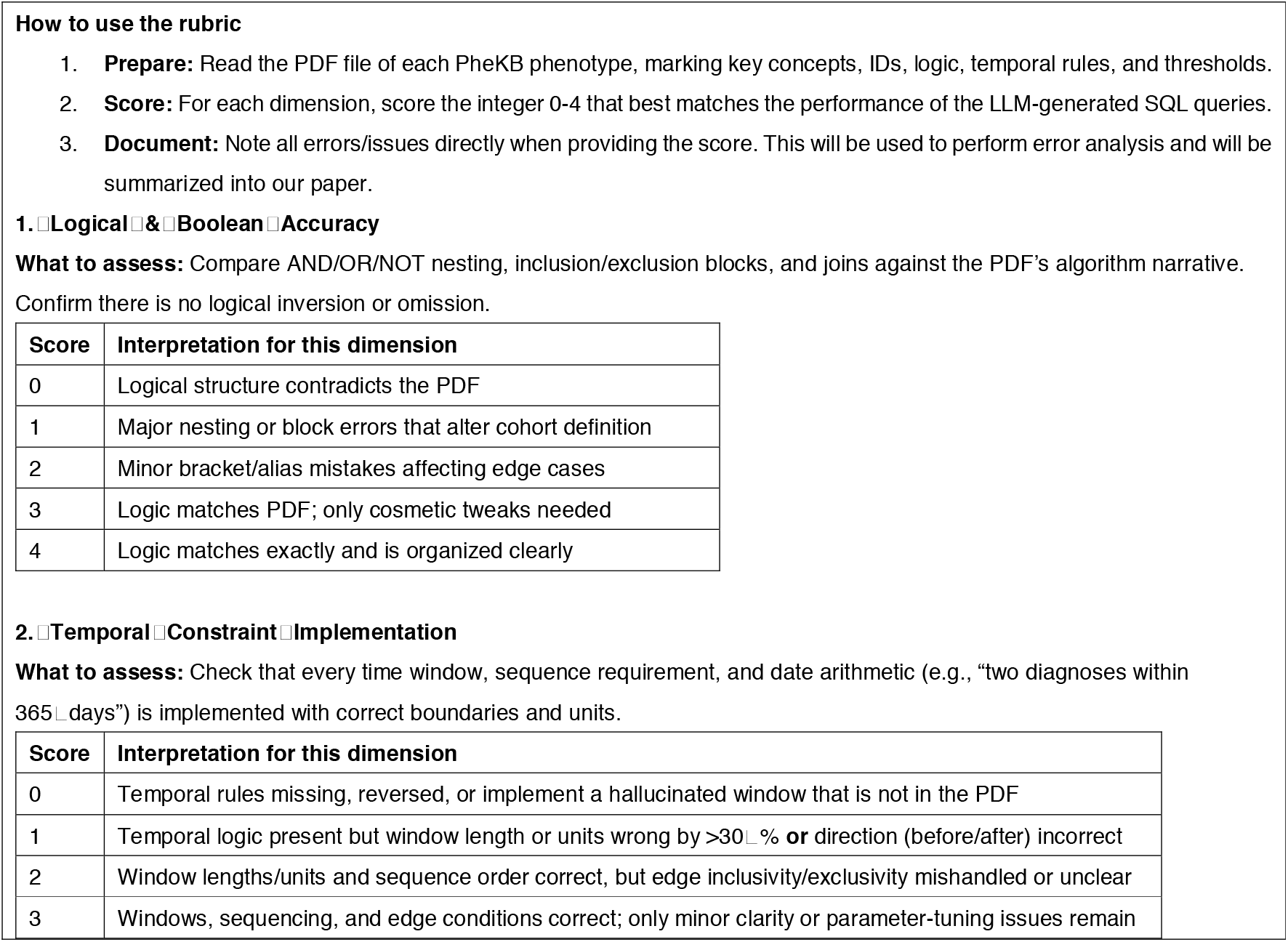
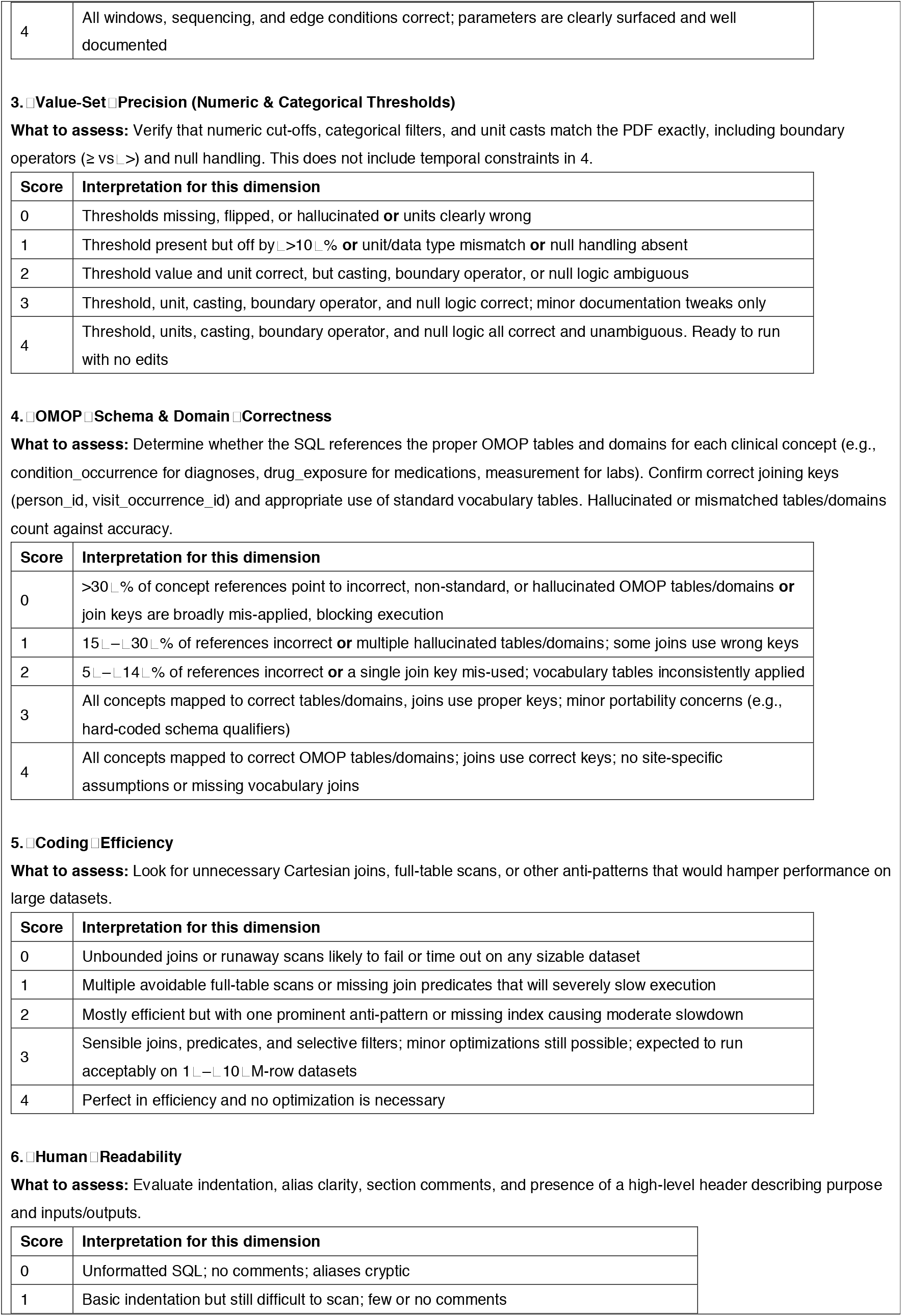
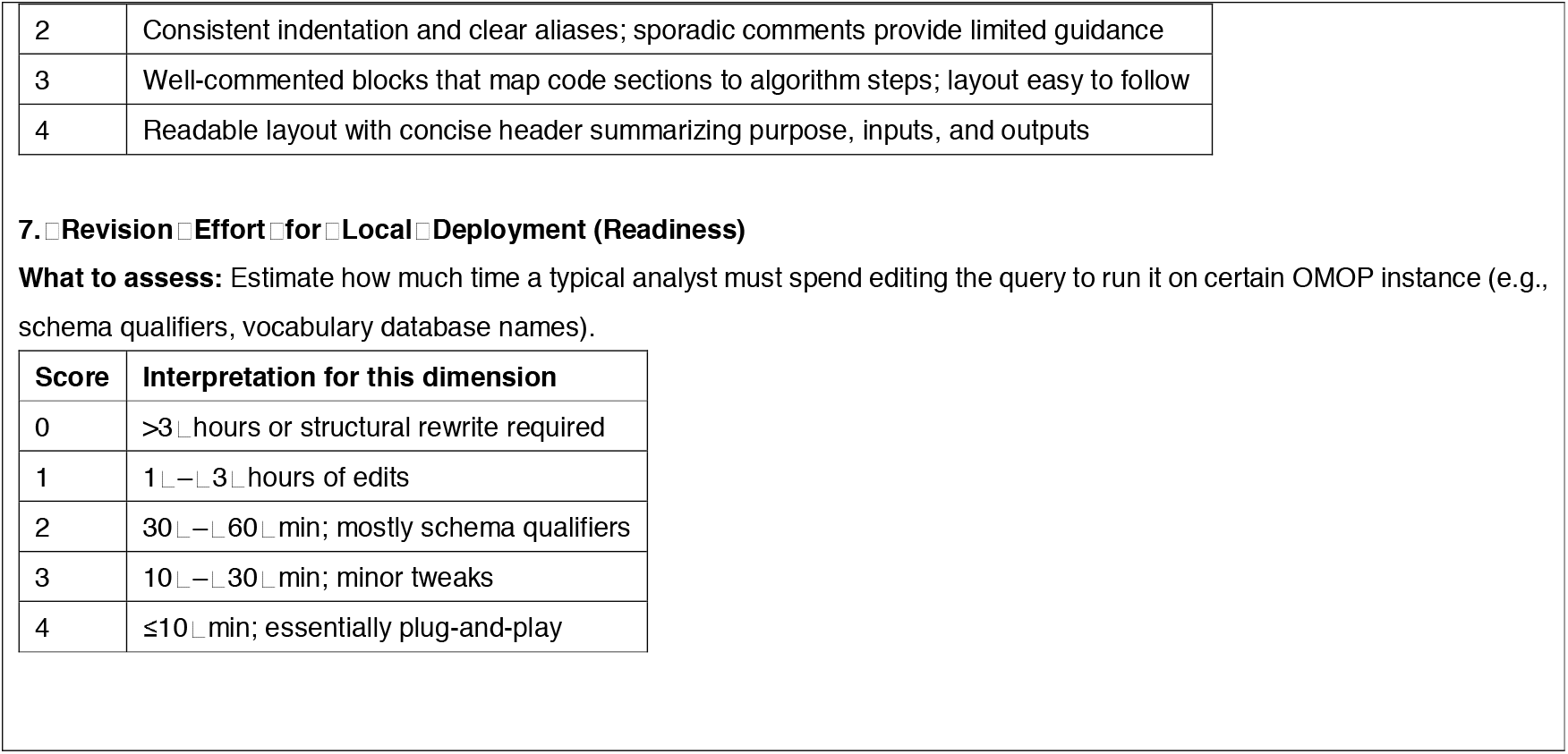
Evaluation criteria used by clinical experts.

**Supplementary Table 3:** Interrater agreement between two clinical experts, measured by Cohen’s kappa for five diseases. ADs: autoimmune disease; AKI: acute kidney injury; FH: familial hypercholesterolemia; MACE: major adverse cardiac events; T2DM: type 2 diabetes mellitus.

| LLM | Phenotyping algorithm | Cohen's kappa (quadratic) | LLM | Phenotyping algorithm | Cohen's kappa (quadratic) |
| --- | --- | --- | --- | --- | --- |
| OpenAI o3 | ADs | 0.9275 | Claude Opus 4.1 | ADs | 0.9356 |
|  | AKI | 0.8094 |  | AKI | 0.8027 |
|  | FH | 0.8222 |  | FH | 0.6316 |
|  | MACE | 0.5714 |  | MACE | 0.9605 |
|  | T2DM | 0.6364 |  | T2DM | 0.9569 |
|  | **Overall** | 0.7974 |  | **Overall** | 0.8977 |

